# PREVALENCE AND TREATMENT OUTCOME OF ANORECTAL MALFORMATION AT MNAZI MMOJA REFERRAL HOSPITAL 2018-2022

**DOI:** 10.1101/2024.09.03.24310510

**Authors:** Ahmad Moshi Chongera, Abdulkarim Hamad Ally, Mwanakhamis Vuai Mussa, Yasinta Edison Mnyaruge, Said A Said, Mwanabaraka Saleh Haji

## Abstract

**BACKGROUND:** Anorectal Malformations (ARM) are congenital defects where the anus and rectum do not form correctly, presenting a range of complexities that often necessitate intricate surgical interventionist global incidence of ARM is approximately 1 in 2000 to 5000 live births. In developing countries like Tanzania, managing ARMs poses significant challenges due to late presentations, shortage of trained pediatric surgeons and inadequate diagnostic facilities. This study aimed to evaluate the prevalence and treatment outcomes of ARM at Mnazi Mmoja Hospital (MMH) over five years 2018 to 2022

**OBJECTIVES:** including analyzing patient demographics (age and sex), the prevalence of ARMs, common types, surgical procedure used and associated complications. A retrospective, descriptive cross-sectional study was conducted using data from structed questionnaire, which were analyzed with SPSS software.

**RESULTS:** revealed that the sex distribution of patients was nearly equal, with ARM cases increasing from 8 (9.9%) in 2018 to 33(40.7%) in 2022. The majority of cases were in Urban Wes (49.9%), and most patients (90.1%) had major ARMs. Surgical treatments included three-stage procedures (65.4%) and one-stage procedures (13.6%), with a postoperative complication rate of 17.2%. survival was high, with 90.1% survival rate and low mortality rate of 9.9%

**CONCLUSION:** ARM remains a prevalent and challenging surgical issue in Zanzibar, constituting 35% 0f pediatric gastrointestinal surgeries. Implementing modern single-stage surgical approaches could potentially improve patient outcomes and reduces complications

## 1.1 BACKGROUND INFORMATION

Anorectal malformations are birth defects in which the anus and rectum (the lower end of the digestive tract) don’t develop properly.

Anorectal malformations (ARMs) are common congenital abnormalities in most parts of the world and their management remains a challenge to surgeons practicing in resource-limited settings (Mfinanga et al., 2018) like in our setting.

Anorectal malformations (ARMs) afflict both sexes, encompass a wide range of illnesses, and affect the distal anus and rectum, as well as the urogenital tracts. Defects can range in severity from those that are small and easily treatable with excellent results to those that are complex and frequently accompanied by multiple anomalies and difficult to control with a poor functional outlook.(Mahmud et al., 2021)

With an incidence of about 1 in 5,000 live births worldwide, ARMs are the one of the most common congenital malformations; however, the incidence seems to be higher in African settings

Males are more likely to experience one anomaly or a mix of abnormalities than females. Up to 70% of cases with ARMs have been documented to be accompanied by other congenital abnormalities. The existence and severity of these related anomalies have a significant impact on the final prognosis and quality of life for children with ARM’s. Patients have the best opportunity for a positive functional outcome with early diagnosis, therapy of related abnormalities, and skillful precise surgical repair (Mfinanga et al., 2018).

In developing countries such as Tanzania, the management of ARMs pose a diagnostic and therapeutic challenge. Late presentation with attendant complications, limited access to trained pediatric surgeons, and the lack of facilities for prompt diagnosis of associated congenital anomalies characterize the poor management of this disease. The conventional approach to surgical repair of most ARMs involves a multi-stage repair, the initial colostomy creation followed by definitive pull-through and eventual colostomy closure (Thapa et al.,2013)

Much research done in most of the world suggests that anorectal malformations are classified as etiological based on the site of the gut involved which are High-type, intermediate-type, and L-type anorectal malformations.

Most pediatric surgeons use the term “low-type” anorectal malformations (ARMs) in cases of rectoperineal or recto vestibular fistula. Low-type ARMs comprise an important subset of the distinct types of anomalies in both sexes, accounting for approximately half of all ARMs(Zheng et al., 2019)

## 2.0 METHODOLOGY

### 2.1 Study design

A hospital-based retrospective, descriptive cross-sectional study.

### 2.2 study area

This study was conducted in the surgical ward at MMH. It is found in the urban west region of Unguja, Zanzibar, Tanzania.

### 2.3 study variables

a) Dependent
  ■ Anorectal malformation
b) Independent
  ■ Age
  ■ Sex
  ■ Type of surgery
  ■ Hospital Stay

### 2.4 Study population

Enrolled pediatric patients with ARMs who undergo surgery between 2018 to 2022.

### 2.5 Inclusive and exclusion criteria

Inclusion

All patient admitted with ARMs and their desired information was available in the hospital records.

Exclusive

All patient with ARM whose information’s was not available in records and those who did not have surgery at MMH

### 2.6 Sample size

81 patients. According to the incidence of unpublished data available at MMH it is roughly estimated that 1 – 2 patients will be diagnosed with ARM in two months making a total of 6 – 12 patients per year and 30 – 60 patients within five years of study

### 2.7 Sampling techniques

information was recorded in a structured questionnaire. Data on each patient was collected and entered into a pretested coded questionnaire prepared for the study

### 3.8 Data analysis and processing

The Statistical Package for the Social Sciences (SPSS) software version 22.0 was used to analyze the data. Tables and chat were used to present the variable outcome

### 3.9 Ethical Consideration

Permission to conduct the study was obtained from the School of Health and Medical Sciences-SUZA as well as MMH through the Pediatric surgical department

Ethical approval was sought from ZAHREC under **REF: NO. ZAHREC/05/ST/MARCH/2023/38**

In this study parents, guardians or informants had to sign an informed written consent for the study on behalf of their children

The study did not interfere with the decision of the attending doctor.

## 3.0 RESULTS

### 1. Socio-demographic characteristics

During the study period, total of 81 patients with major congenital malformations were managed at Mnazi Moja Hospital. Forty (49.4%) were males and 41(50.6%) were females giving a male to-female ratio of nearly 1: 1. (figure 1)

**Figure 1:**
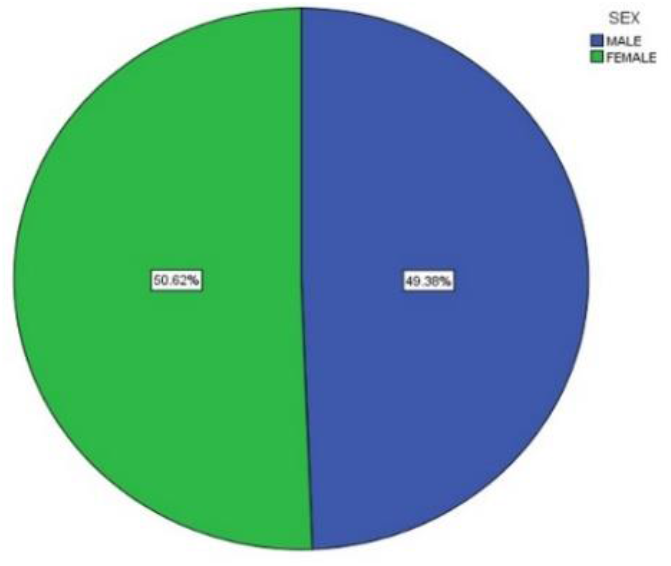
Sex distribution of the patient with anal rectal malformations admitted at Mnazi Mmoja Hospital.

### 2. Age distribution of the patient with anal rectal malformations admitted at Mnazi Mmoja Hospital

Of which 44 children (54.3%) were aged between 0-2 days, 28 children (34.6%) were aged between 3-5 days and the rest 9 were aged between >5 days (11.1%) as shown in figure 2.

**Figure 2:**
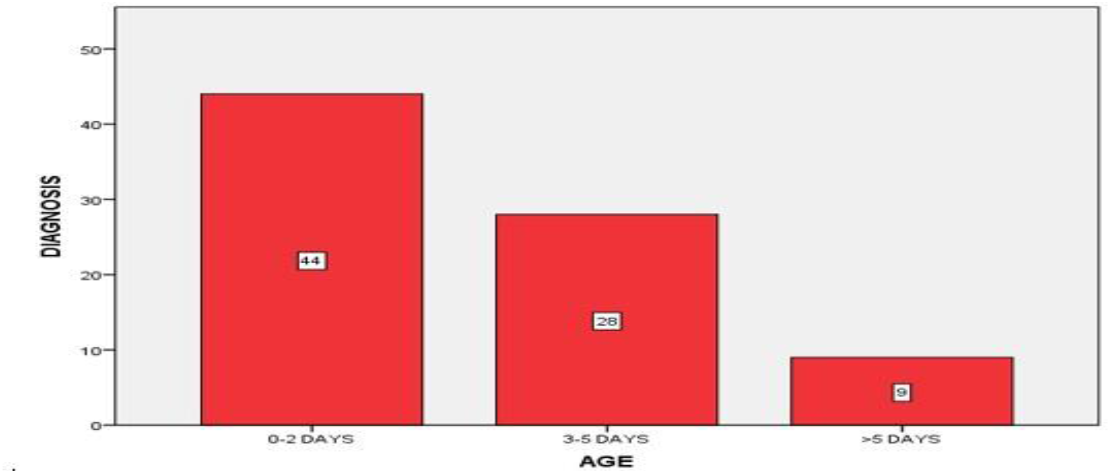
Age distribution of the patient with anal rectal malformations admitted at Mnazi Mmoja Hospital.

The age at diagnosis ranged between a few hours after birth to more than 5 days with a median age of 1 day.

### 3. Admission year for the patient with anal rectal malformations admitted at Mnazi Moja Hospital

The distribution of the number of patients with their respective years of treatment is shown in Table 1

**Table 1:** Admission year for the patient with anal rectal malformations admitted at Mnazi Moja Hospital (MMH)

| ADMISSION YEAR | FREQUENCY | PERCENT |
| --- | --- | --- |
| 2018 | 8 | 9.9 |
| 2019 | 5 | 6.2 |
| 2020 | 17 | 21.0 |
| 2021 | 18 | 22.2 |
| 2022 | 33 | 40.7 |
| Total | 81 | 100.0 |

### 4. The geographical distribution of patients admitted during the study period is summarized in figure 3 and Table 2

**Table 2:** Geographical distribution of patient admission with respective year for the patient with anal rectal malformations admitted at MMH.

|  |  |  | ADMISSION YEAR |  |  |  |  | Total |
| --- | --- | --- | --- | --- | --- | --- | --- | --- |
|  |  |  | 2018 | 2019 | 2020 | 2021 | 2022 |  |
| REPORTED AREA | PEMBA | Count | 2 | 2 | 3 | 5 | 6 | 18 |
|  |  | % of Total | 2.5% | 2.5% | 3.7% | 6.2% | 7.4% | 22.2% |
|  | SOUTH UNGUJA | Count | 0 | 1 | 2 | 0 | 6 | 9 |
|  |  | % of Total | 0.0% | 1.2% | 2.5% | 0.0% | 7.4% | 11.1% |
|  | NORTH UNGUJA | Count | 2 | 2 | 5 | 3 | 2 | 14 |
|  |  | % of Total | 2.5% | 2.5% | 6.2% | 3.7% | 2.5% | 17.3% |
|  | URBAN WEST | Count | 4 | 0 | 7 | 10 | 19 | 40 |
|  |  | % of Total | 4.9% | 0.0% | 8.6% | 12.3% | 23.5% | 49.4% |
| Total |  | Count | 8 | 5 | 17 | 18 | 33 | 81 |
|  |  | % of Total | 9.9% | 6.2% | 21.0% | 22.2% | 40.7% | 100.0% |

### 5. Complains distribution of the patient with anal rectal malformations admitted at Mnazi Mmoja Hospital

The majority of patients 68 (84%) had imperforated anus without fistula and 8 (9.9%) had associated fistula (Figure 4).

**Figure 3:**
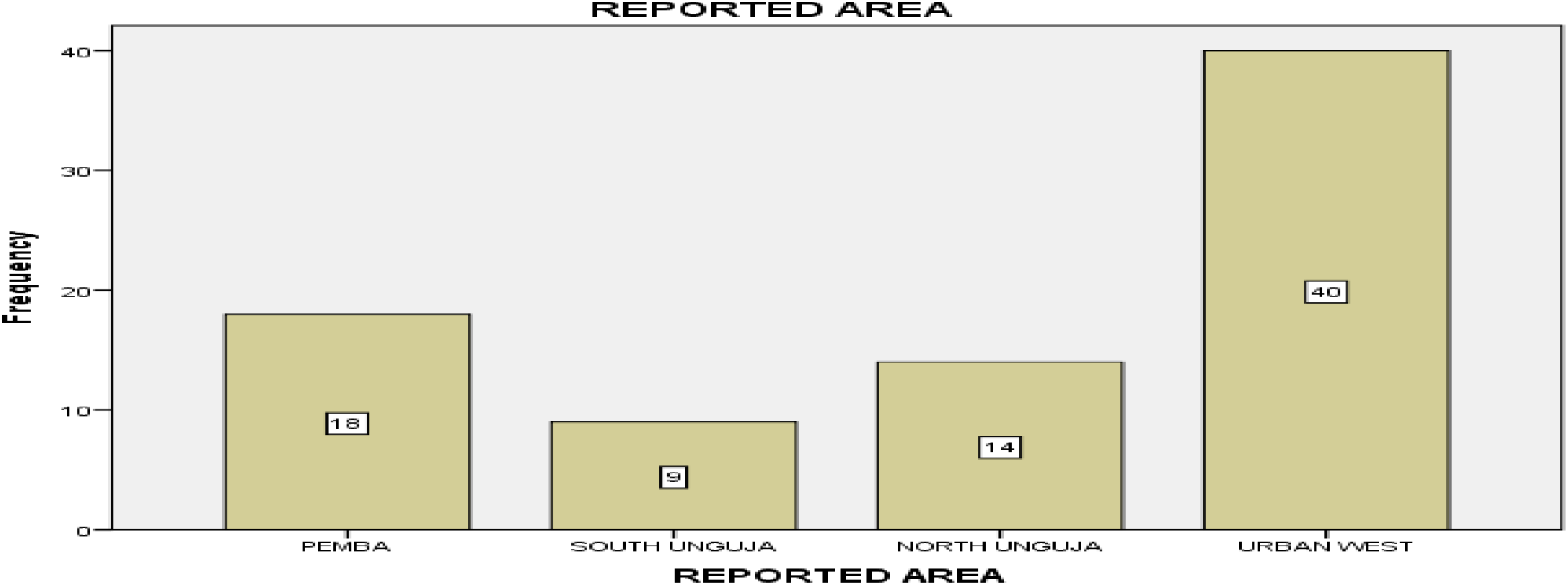
distribution area for patient with anal rectal malformations admitted at MMH

**Figure 4:**
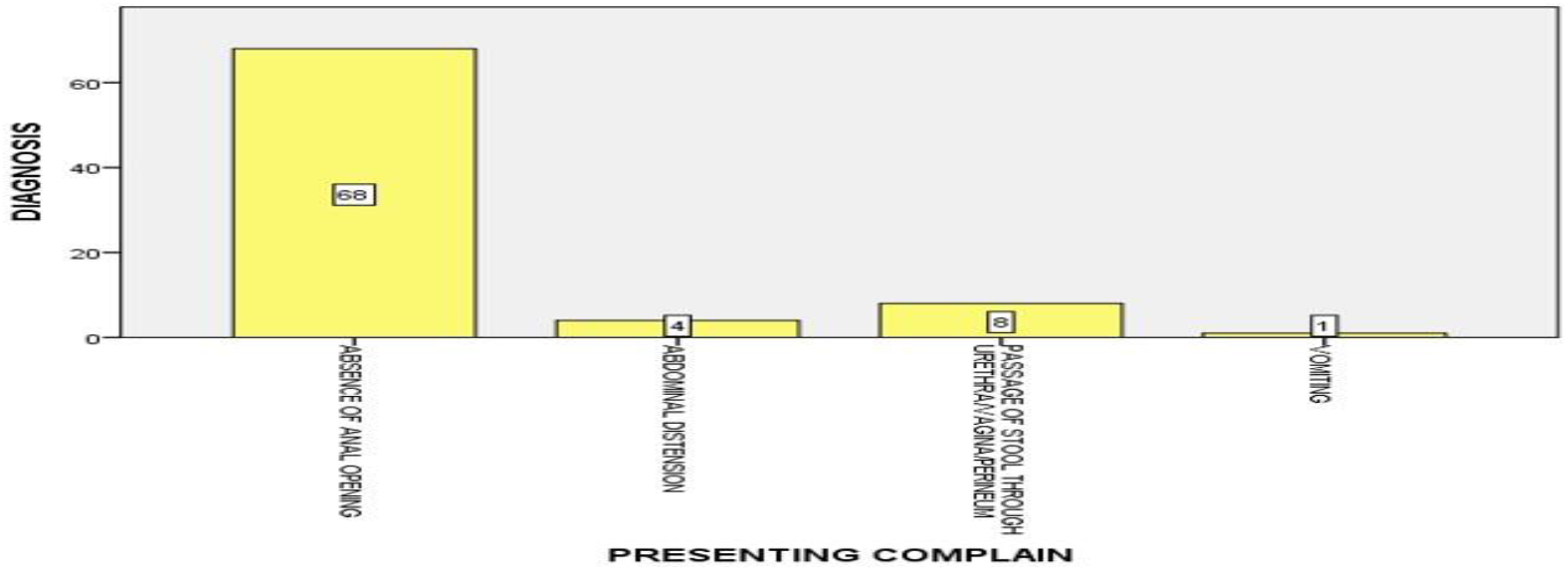
Complains distribution of the patient with anal rectal malformations admitted at Mnazi Mmoja Hospital.

**Figure 5:**
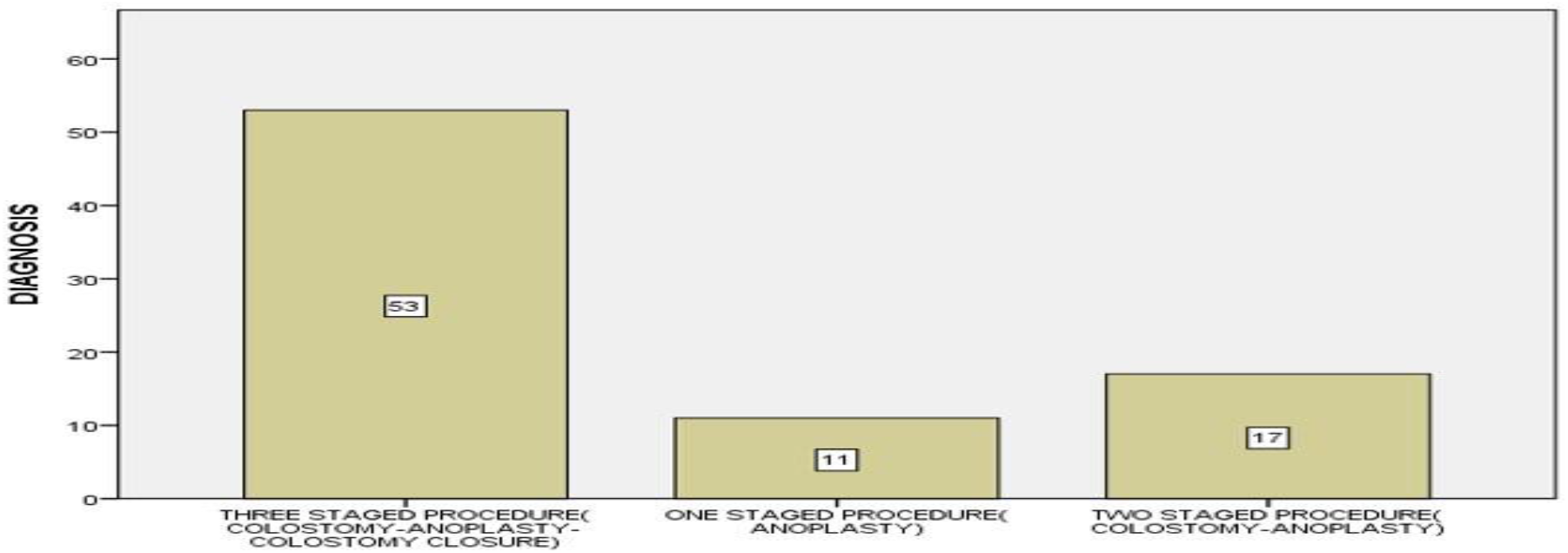
Surgical management for the patient with anal rectal malformations admitted at MMH

### 6. Time for the first diagnosis for the patient with anal rectal malformations admitted at MMH

Majority of patients were admitted soon after birth 30(37%) as shown in Table 3

**Table 3:** Time for the first diagnosis the patient with anal rectal malformations admitted at MMH.

| TIME FOR DIAGNOSIS | FREQUENCY | PERCENT |
| --- | --- | --- |
| SOON AFTER BIRTH | 30 | 37.0 |
| < 24 HRS | 27 | 33.3 |
| >24 HRS | 24 | 29.6 |
| WEEK/MONTH | 0 | 0 |
| Total | 81 | 100.0 |

### 7. Diagnosis for the patient with anal rectal malformations admitted at MMH

According to Krickenbeck’s classification of ARMs (Holschneider et al., 2005), the majority of patients, 73 (90.1%) had major clinical type of ARMs and 8 (9.9%) had minor clinical type (Table 4)

**Table 4:** Distribution of patients according to Krickenbeck classification of ARMs.

|  |  |  |  |  |
| --- | --- | --- | --- | --- |
| 1. | CLOACAE | 6 (7.4%) |  | 6 (7.4%) |
| 2. | RECTO-VESTIBULAR FISTULA | 10 (12.3%) |  | 10 (12.3%) |
| 3. | RECTO-VESICLE FISTULA | 0 (0%) |  | 0 (0%) |
| 4. | IMPERFORATED ANUS (ARM WITHOUT FISTULA) | 57 (70.4%) |  | 57 (70.4%) |
| 5. | ANAL STENOSIS | 0 (0%) |  | 0 (0%) |
| 6. | RECTO-URETHRAL FISTULA | 0 (0%) |  | 0 (0%) |
| 7. | PERINEAL FISTULA | 0 (0%) |  | 0 (0%) |
| 8. | RECTO-VAGINAL FISTULA |  | 8 (9.9%) | 8 (9.9%) |
| 9. | H-TYPE FISTULA |  | 0 (0%) | 0 (0%) |
| 10. | RECTAL ATRESIA |  | 0 (0%) | 0 (0%) |
|  | TOTAL | 73 (90.1%) | 8 (9.9%) | 81 (100%) |

### 8. Surgical management for the patient with anal rectal malformations admitted at MMH

The majority of the patients 70 (86.4%) were planned for three-stage surgery whereas 53(65.4%) had completed surgical intervention. 17(21.0%) was not yet completed and the remaining 11(13.6%) had one stage procedure.

### 9. Treatment outcome for the patient with anal rectal malformations admitted at MMH

Of the 81 patients, 14 developed postoperative complications giving a complication rate of 17.2%. Colostomy prolapse 5 (6.2%) occurred most frequently. Different reported complications are shown in the (Table 5)

**Table 5:**
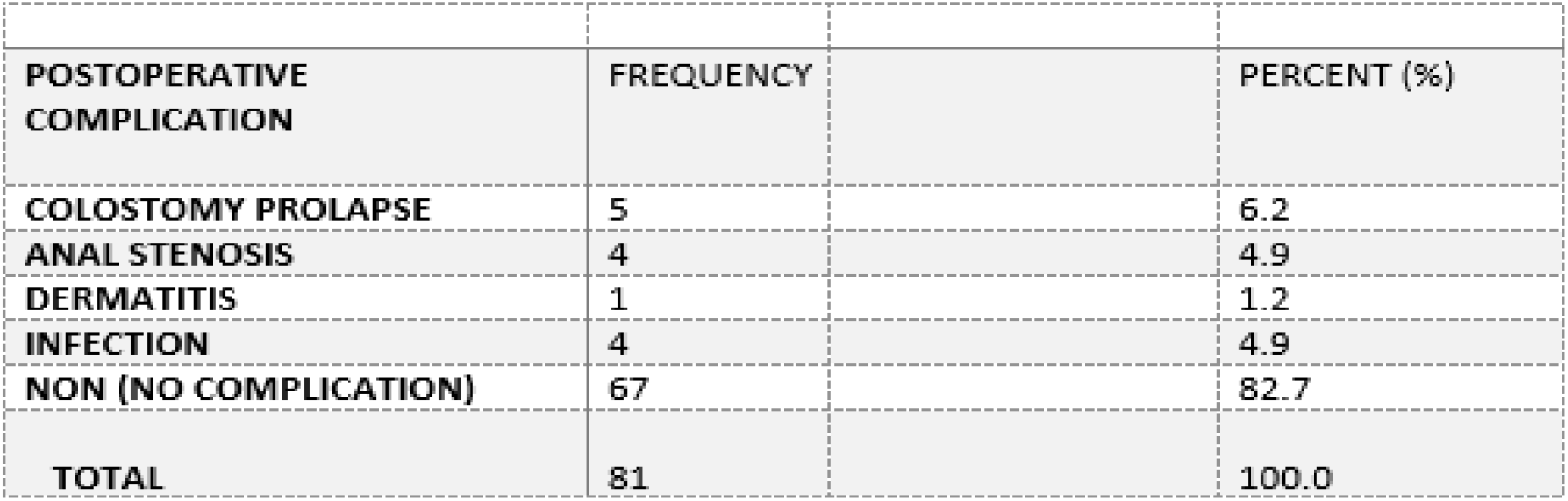
Distribution of complications postoperative for the patient with anal rectal malformations admitted at MMH.

The majority of patients 73 (90.1%) were discharged with satisfactory health status giving an overall survival rate of 90.1% but the remaining patient 8 (9.9%) died after the operation making the mortality rate to be 9.9% (Figure 6)

**Figure 6:**
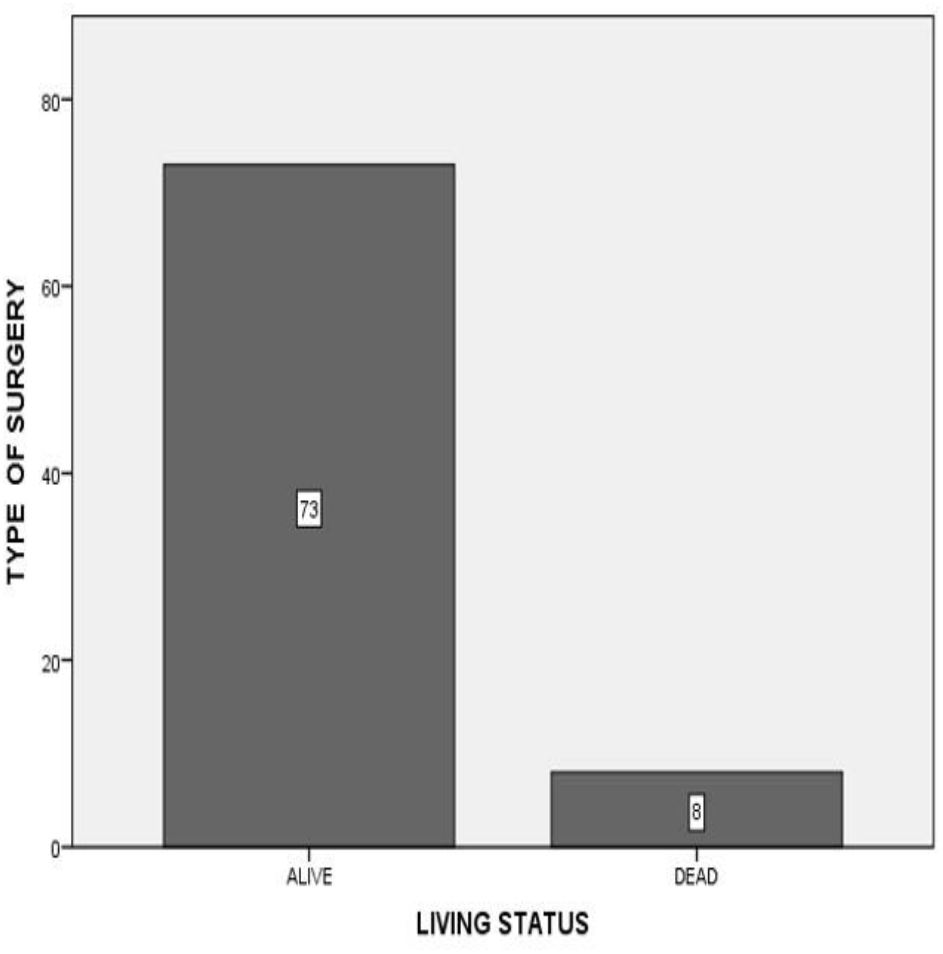
Treatment outcome for the patient with anal rectal malformations admitted at MMH

### 10. Distribution of Hospitalization for the patient with anal rectal malformations admitted at MMH

The majority 52 (64.20%) of our patients were postoperatively hospitalized for less 10 days with a minimum of a few hours due to post-operative death and more than 15 days. The mean hospital stay was 12.5 days (Figure 7)

**Figure 7:**
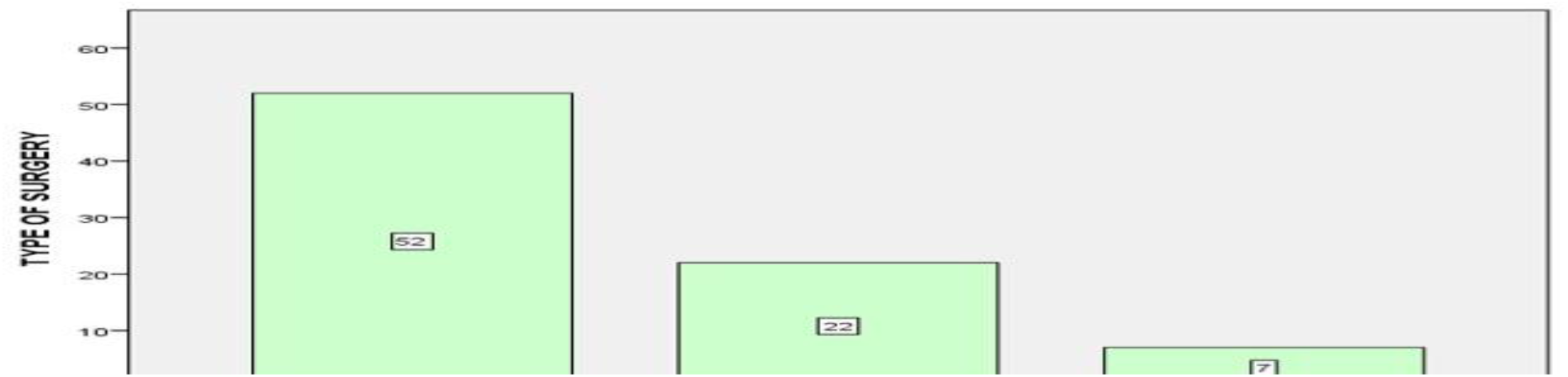
Distribution of Hospitalization for the patient with anal rectal malformations admitted at MMH

## 4.0 Discussion

Anorectal malformation is one of the common pediatric surgical problems seen in Zanzibar Island. In a five-year, period between 2018 and 2022 total of 81 children were treated for ARMs at Mnazi Mmoja referral hospital. In this study, ARMs affect equally for both sexes making a male-to-female ratio of 1: 1. This is contrary to the study done elsewhere where there was male predominance (in Rwanda M. Makanga et al). The findings of this study of an equal ratio of males to females are supported by the study of Bhargava et al,2006. We could not find in the literature the reasons for these gender differences and this warrants further investigation. (Mfinanga et al).

Of 81 patient’s majority 44 % were diagnosed within 2 days of life although delayed diagnosis was found in 28 % at 3 – 5 days after birth and 9 % were diagnosed after the age of 5 days. Similar finding reported by 23.68% of the diagnosis was made beyond the neonatal period (send statovci. Salih graccevci, Murat, Murat Belisha et al, 2015) late diagnosis is contributed to the presence of fistula and ectopic anus in majority of patients where the meconium pass and obscure the presence of ARMs.

The commonest type of ARMs was imperforate anus without fistula which for 57% of patients, Vestibular anus10%, cloaca 6% and rectovaginal fistula 8%. Similar observation was reported by kuradusenge where imperforate anus without fistula was the predominant type seen in 31–42% of boys with ARM in Kenya (Kuradusenge et al and kigo et al). this type of ARMs necessitate early presentation to hospital due to associated signs and symptoms of intestinal obstruction furthermore the passage of meconium even from the associated fistula persuade the nurses and care taker that the child is normal without conducting genital examination.

During the study period approach of ARMs treatment was 3 stage procedure as routine in majority of children 70 (86.4%). Unlike recent approach elsewhere where the primary PSARP is the preferred treatment option (Peña & Devries, (Bischoff et al., 2013)1982; Peña, 1985; Ratan et al., 2004; Osilo et al., 2014). This is because limited gastroenterological pediatrics surgeons in the region where initial consultation is made by general surgeons who perform colostomies on an emergency basis.

In the majority of the patients, 68 (84%) diagnosed with ARMs their major complaint was the absence of an anus. A similar result was obtained by Makanga et al absence of an anus in 39.1% of cases. This is mail contributed by the fact that the absence of an anus especially when no passage of meconium threats caregiver and sends the children for medical care.

The majority of the patients 37 % admitted at MMH in a pediatrics surgical ward with ARM was diagnosed soon after delivery and there was no patient diagnosed more than a week in contrast with (Birabwa-Male, 2004; Makanga et al., 2007) where majority of ARM patients reported to the hospital late in acute intestinal obstruction which required emergency surgical intervention. This observation has an implication on good accessibility to healthcare facilities in Zanzibar in terms of short distance to reach health facility and also free services for all citizens.

In this study the majority of the ARM patients 52 (64.20%) post operatively were hospitalized for less than 10 days among 52 (64.20%) this is in contrary with the study done elsewhere where less duration is used by the patient after surgery 3 to 5 days. (Qureshi et al., 2022).

Of the 81 patients. Fourteen patients reported to developed postoperative complications giving a complication rate of 17.2%. Major complication was stoma related similar to the findings obtained by Birabwa Male D. anal stenosis of neo anus and colostomy prolapse were major concern among the patients in many centers. In this study we observed colostomy prolapse 6.2% anal stenosis and stoma infection 4.9% with stoma dermatitis1.2%. Much lower complications rate 14% was reported in other study with Makanga et al. these might be contributed to single stage procedures which is the main approach is these center which minimize the delay which occurs in between the treatment stages.

Of 81 patient 73 were alive and was discharged home with no complications, and 8 (9.9%) died of surgery related complications. In other study higher mortality rate 17.6% was reported (Mantho P1,2, Essola B2, Asongwe M3, Tumameu T4, Bitchoka ER1, Kamguep T5, Azo’o Mfou’ou F6, Mefire CA3,6, Engbang JP1,2). Early intervention and accessibility to proper surgical care was the reason for low mortality rate in our setting.

Following definitive surgery, complications are reported among one third of the patients (Table 5) although actual figures may be higher in some series as patients may be lost to follow up and some of those may die (Taiwo A Lawal)

## 5.0 CONCLUSION

Zanzibar as in other part of the world the ARMs is common surgical problems causing considerable morbidity and mortality. For reducing diagnosis delay a careful, comprehensive clinical examination of genitalia in all new born in PHCU+ and hospital level both government and private stake holder effort is required.

Also, parent awareness and reporting will help home delivery women.

## 6.0 RECOMMENDATIONS

More studies in ARMs and associated congenital anomalies and risk factors is required in order to improve treatment out come

To improve skills of general surgeons in pediatric surgeries due to scarcity of Pediatric surgeons

Health education to improve awareness of health care providers and community on routine examination of new born genitalia soon after delivery

## Supporting information

This is table of data for patient used for analysis

This are sample of questionnaire questions and code used which are corresponding to number filled in excel for patient used for analysis

## Data Availability

All data produced in the present study are available upon reasonable request to the authors

