## Supplementary material for "PREVALENCE AND TREATMENT OUTCOME OF ANORECTAL MALFORMATION AT MNAZI MMOJA REFERRAL HOSPITAL 2018-2022": This are sample of questionnaire questions and code used which are corresponding to number filled in excel for patient used for analysis

### **QUESSIONARE FORM**

**DEFINATION OF CODE USED DURING ANALYSIS**

**A) ADMISION YEAR**

1. 2018
2. 2019
3. 2020
4. 2021
5. 2022

**B) REPORTED AREA**

1. PEMBA

2. SOUTH UNGUJA

3. NORTH UNGUJA

4. URBAN WEST

**C) AGE**

1. 0-2 DAYS
2. 3-5 DAYS
3. >5 DAYS

**D)SEX**

1. MALE
2. FEMALE

**E) PRESENTING COMPLAINS**

1. ABSENCE OF ANAL OPENING
2. ABDOMINAL DISTENTION
3. PASSAGE OF STOOL THROUGH URETHRAL
4. VOMITING

**F) DIAGNOSIS**

1. IMPERFORATED ANUS
2. VRSTIBULAR ANUS
3. CLOACAE
4. RECTAL VAGINAL FISTULA

**G) TIME OF FIRST DIAGNOSIS**

1. SOON AFTER BIRTH
2. < 24 HOURS
3. > 24 HOURS
4. WEEK/MONTH

**H) PLACE OF DIAGNOSIS**

1. DELIVERY CENTER
2. HOME
3. HOSPITAL

**I)TYPES OF SURGERTY FOR MANAGEMENT**

1. THREE STAGED PROCEDURES
2. ONE STAGED PROCEDURES
3. TWO STAGED PROCEDURES

**J) COMPLICATION**

1. COLOSTOMY PROLAPSE
2. ANAL STENOSIS
3. DERMATITIS
4. COLOSTOMY RETRACTION
5. INFECTION
6. NONE

**K) LIVING STATUS**

0.ALIVE

1. DEAD

**L) HOSPITAL STAY**

1. < 10 DAYS

2. 10-15 DAYS

3. > 15 DAYS

**M) ANORECTAL MAFORMATION TYPES**

1. MAJOR

2.MINOR
